# Fecal metaproteomics enables functional characterization of remission in patients with inflammatory bowel disease

**DOI:** 10.1101/2024.07.02.24309587

**Authors:** Maximilian Wolf, Julian Lange, Dirk Benndorf, Lina Welz, Susanna Nikolaus, Laura Sievers, Florian Tran, Kay Schallert, Patrick Hellwig, Stefan Schreiber, Matthias Gunzer, Philip Rosenstiel, Udo Reichl, Konrad Aden, Robert Heyer

## Abstract

**Background:** The gut microbiome is an important contributor to the development and the course of inflammatory bowel disease (IBD). While changes in the gut microbiome composition were observed in response to IBD therapy using biologics, studies elucidating human and microbial proteins and pathways in dependence on therapy success are sparse.

**Methods:** Fecal samples of a cohort of IBD patients were collected before and after 14 weeks of treatment with three different biologics. Clinical disease activity scores were used to determine the clinical response and remission. Fecal metaproteomes of remitting patients (n=12) and of non-remitting patients (n=12) were compared before treatment and changes within both groups were assessed over sampling time to identify functional changes and potential human and microbial biomarkers.

**Results:** The abundance of proteins associated with the intestinal barrier, neutrophilic granulocytes, and immunoglobulins significantly decreased in remitting patients. In contrast, an increase of those proteins was observed in non-remitting patients. There were significant changes in pathways of microbial metabolism in samples from patients with remission after therapy. This included, for example, an increased abundance of proteins from butyrate fermentation. Finally, new potential biomarkers for the prediction and monitoring of therapy success could be identified, e.g. human lysosome-associated membrane glycoprotein 1, a cytotoxicity marker, or microbial anthranilate synthase component 2, a part of the tryptophan metabolism.

**Conclusions:** Distinct changes of proteins related to gut inflammation and gut microbiome metabolism showed whether IBD remission was achieved or not. This suggests that metaproteomics could be a useful tool for monitoring remission in IBD therapies.

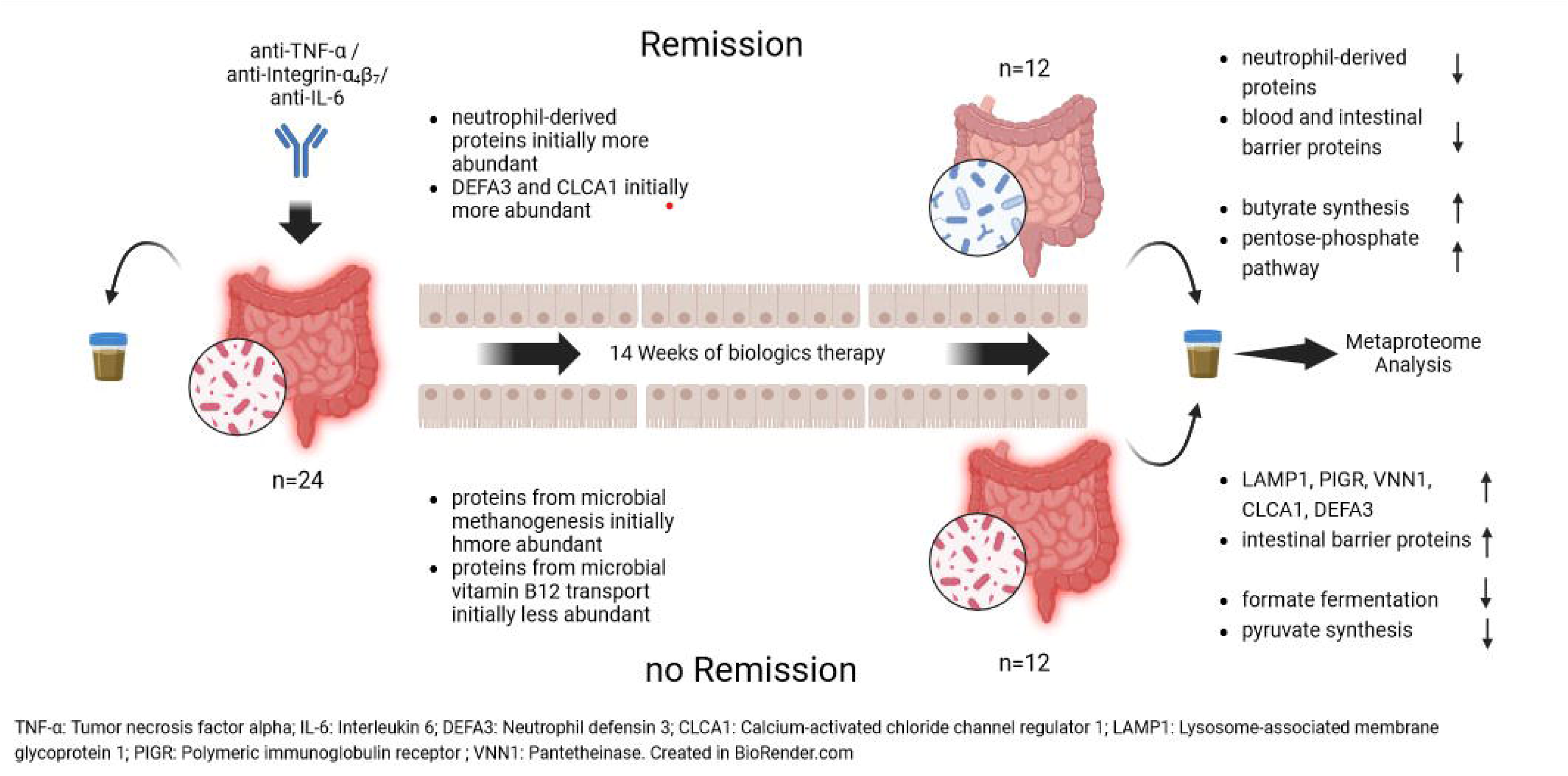

## 1. Introduction

Crohn’s disease (CD) and ulcerative colitis (UC), summarized under the term inflammatory bowel diseases (IBD), are chronic, recurrent inflammations of the intestinal tract that place enormous physical and psychosocial burdens on the patient [1, 2]. IBD has been shown to be linked to the patient’s genetics and environmental factors. However, the patient’s intestinal microbiome also plays a special role in the etiology [3–6]. The permanent exposure of the intestinal epithelium to diverse microbial antigens requires a fine orchestration of the immune response in the case of pathogen infection, while still exhibiting a tolerance of commensal microorganisms [7]. This is ensured in the intestine of healthy individuals by an intact epithelial barrier as well as a broad spectrum of immunologically active substances, such as cytokines or adhesion molecules. In patients with IBD, the inflammatory response is inappropriate and the epithelial barrier is damaged, which can lead to infiltration of the intestinal tissue with microorganisms and consequent inflammatory cascades [8].

Various therapeutic approaches such as drug, surgery, nutritional intervention, and fecal transplantation are used to treat this inflammation and to achieve remission, a medical condition characterized by decreasing severity of disease symptoms and mucosal healing [9]. Typical drugs are small-molecule drugs and biologics. Small molecule drugs, such as thiopurines, aminosalicylates, or steroids, are inexpensive, non-immunogenic molecules, but are characterized by a short half-life and low selectivity and efficacy [10, 11]. In contrast, biologics are large complex molecules, in the form of monoclonal antibodies or antibody-drug-conjugates, which specifically bind to certain molecules involved in IBD pathogenesis, particularly those that interfere with immune cell communication or migration [12]. Although monoclonal antibodies have a higher half-life, effectiveness, and selectivity, antibodies can be recognized as antigens by the patient’s immune system and thus their long-term effectiveness can be severely limited. In addition to the immunogenicity of the antibodies, other factors contribute to widely varying efficacies of treatments in different patients, such as genetic polymorphisms or metabolic potential of the patient’s gut microbiome [13–15].

Despite increasing efforts and the introduction of novel targeted therapies in the treatment of IBD (anti-TNF, anti-a_4_b_7_ integrin, anti-IL12/23, anti-IL23, JAK inhibitors, S1P modulators) the overall long-term disease control is still considerably low at approximately 40% [16].

The early assessment of biomarkers that are associated with a favorable or unfavorable treatment response to biologic therapies might therefore help to guide early treatment decision (e.g. switch treatment class). Huge efforts have been undertaken to investigate the role of the intestinal microbiome as a potential driver of treatment response in IBD. In this context most studies have initially focused on analysis species diversity using 16S analysis, whilst lately metabolic properties (in-silico metabolic modelling [17], fecal metabolomics [18]) have been the focus of investigation.

Although the metaproteome reflects the actual proteins expressed in the microbiome and the human proteins in the stool sample and might show a stronger association with the inflammation state as gen-based methods [19], there are only very few publications on longitudinal studies of IBD patients using metaproteomic methods compared to a multitude of metagenomic studies [20–24]. Therefore, this study aimed to use metaproteomic approaches to investigate the host and microbial metabolism as well as the abundance of proteins representing the host’s immune response, to determine differences between patients who achieved remission with biologics therapy and those who did not.

## 2. Material and Methods

### Patients and sampling

In the course of this study, the 26 patients with IBD, including 13 patients diagnosed with CD and 13 patients diagnosed with UC, were treated with three different therapeutic agents (Infliximab: n=7, Vedolizumab: n=9, Olamkizept n=10) at the University Hospital Schleswig Holstein, as described before [13, 25]. At baseline and after 14 weeks of treatment, C-reactive protein (CRP) and Interleukin 6 (IL-6) in the blood, calprotectin and blood leukocytes in the stool of the patients were measured by ELISA. Disease intensity was characterized using the MAYO score for UC and the Harvey-Bradshaw index (HBI) for CD patients. The participants provided paired stool samples at baseline and after 14 weeks of therapy. To inactivate potential pathogenic material during sample processing, 200 µL of 1% Sodium docecyl sulfate (SDS) solution were added to 200 µg of the stool samples and this solution was heated to 99°C for 10 minutes.

### Metaproteomic analysis

Metaproteomic analysis was performed according to a previously established workflow [26, 27]. The proteins of the solutions were extracted using phenol in a ball mill. The protein concentration was then determined using the Amido Black assay. Subsequently, 10 µg protein of the samples were tryptically digested (enzyme-substrate ratio 1:100) on a filter according to the filter-aided sample preparation (FASP) protocol [28]. LC/MS-MS measurements were performed using an UltiMate 3000 RSLCnano chromatography system (Thermo Fisher Scientific, Bremen, Germany) with a C18 reversed phase pre-column (Acclaim PepMap100 C18, Thermo Fisher Scientific, Bremen, Germany) and a C18 reversed-phase separation column (Acclaim PepMap100 C18, Thermo Fisher Scientific, Bremen, Germany) coupled to a timsTOF mass spectrometer (Bruker Daltonik GmbH, Bremen, Germany). Further details are summarised in Supplementary Note 1. Compass DataAnalysis 5.1 software (Bruker Daltonik GmbH, Bremen, Germany) was used to process and analyze the mass spectra. Using the search engine Mascot™ 2.6 (Matrix Science, London, Great Britain) and the following parameters: enzyme trypsin, one missed cleavage, monoisotopic mass, carbamidomethyl as fixed and oxidation as variable modification, ±0.02 Da precursor and ±0.02 Da MS/MS fragment tolerance and a false discovery rate of 1 %, the peptides were annotated with the entries of UniProtKB/SwissProt (16/01/2019) and a metagenome database published by Qin et al. [29], used in a previous IBD study [30]. Identifications of redundant homologous proteins generated by these searches were combined into protein groups (hereafter referred to as metaproteins) based on shared peptides using MetaProteomAnalyzer, version 3.4. Metaproteins without functional or taxonomic annotation were assigned using the BLAST and the UniProtKB/SwissProt database. Blast hits with an e-value <10^-4^ were used for annotation according to the lowest common ancestor rule. Metaproteins with less than 10 spectra were excluded from the following analysis. Subsequently, the spectral counts of the protein groups were normalized to the total spectral count of the respective sample. Finally, a result matrix additionally consisting of taxonomic annotation, enzyme commission (E.C.) number, Kyoto Encyclopedia of Genes and Genomes (KEGG) orthologies, UniProtKB reference cluster, and UniProt keywords was generated, which was used for the analysis. The MS files are available under accession number PXD053257 in the PRIDE Archive.

### Orthogonal validation of Lysosomal-associated membrane protein 1 (LAMP1) with independent validation cohort using ELISA

For validation of biomarker discovery from metaproteomics experiments, fecale aspirates of n = 58 patients with UC aged 20-80 years were sampled between 2021 and 2023 at the First Medical Department of University Hospital Schleswig-Holstein, Campus Kiel, Germany. All patients provided written informed consent and biomaterial sampling from patients was approved by the local ethics committee of Kiel University (Vote#: B231/98). Fecale aspirates were sampled during colonoscopy by flushing the distal colon with water followed by aspiration of the fluids. Patients were recruited as a broad cross-sectional cohort at different stages of disease activity and displayed an equal distribution of endoscopic Mayo (eMayo) scores (0 – 3) and complete Mayo scores (0 – 11). Remission was defined as an endoscopic Mayo Score of ≤ 1 and a complete Mayo score of ≤ 2. Samples were kept frozen at -80 degrees and measured by ELISA according to the manufacturer (LAMP1: Abcam, ab277464) at the First Medical Department of University Hospital Innsbruck, Austria. Measurements below the detection level were excluded from the final analysis.

### Statistical analysis

For statistical analysis, R-statistics (version 1.3.1093) was used. Unpaired samples (samples at baseline or week 14 from patients with and without remission) were analyzed applying Wilcoxon rank sum test, paired samples (samples at baseline and week 14 from patients with remission, samples at baseline and week 14 from patients without remission) were analyzed applying Wilcoxon signed rank test using the method “wilcox.test”. The visualization with violin plots was realized using the libraries “ggplot2” and “ggstatsplot”. Volcano plots were created using Python (version 3.8.13) and the libraries “pandas”, “numpy”, and “matplotlib”.

## 3. Results

### Characteristics of the study cohort

In total, 26 patients provided fecal samples before and after 14 weeks of biologics therapy. The therapy with one of three biologics (Infliximab: n=7, Vedolizumab: n=9, Olamkizept n=10) led to an overall significant decrease in clinical disease scores (HBI: from 9.91 ± 7.32 to 4.18 ± 4.78, p-Value 0.042)(Mayo score: from 7.15 ± 2.48 to 3.00 ± 2.99, p-value < 0.001)(Figure 1). As remission was characterized as a Mayo score ≤2 respectively an HBI≤4 and a concentration of serum CRP ≤5 mg/L, 12 out of 26 patients reached by definition a remitting state of the disease, 12 did not achieve remission and 2 patients were in remission during the whole therapy. 19 patients showed an overall response to the therapy. In line with the MAYO and HBI score, the concentration of Calprotectin decreased significantly by 77%, CRP by 73% and blood leucocytes by 33% across all patients. The concentration of IL-6 was also reduced after therapy by 27%, but this difference was not statistically significant (Figure 1).

**Figure 1.**
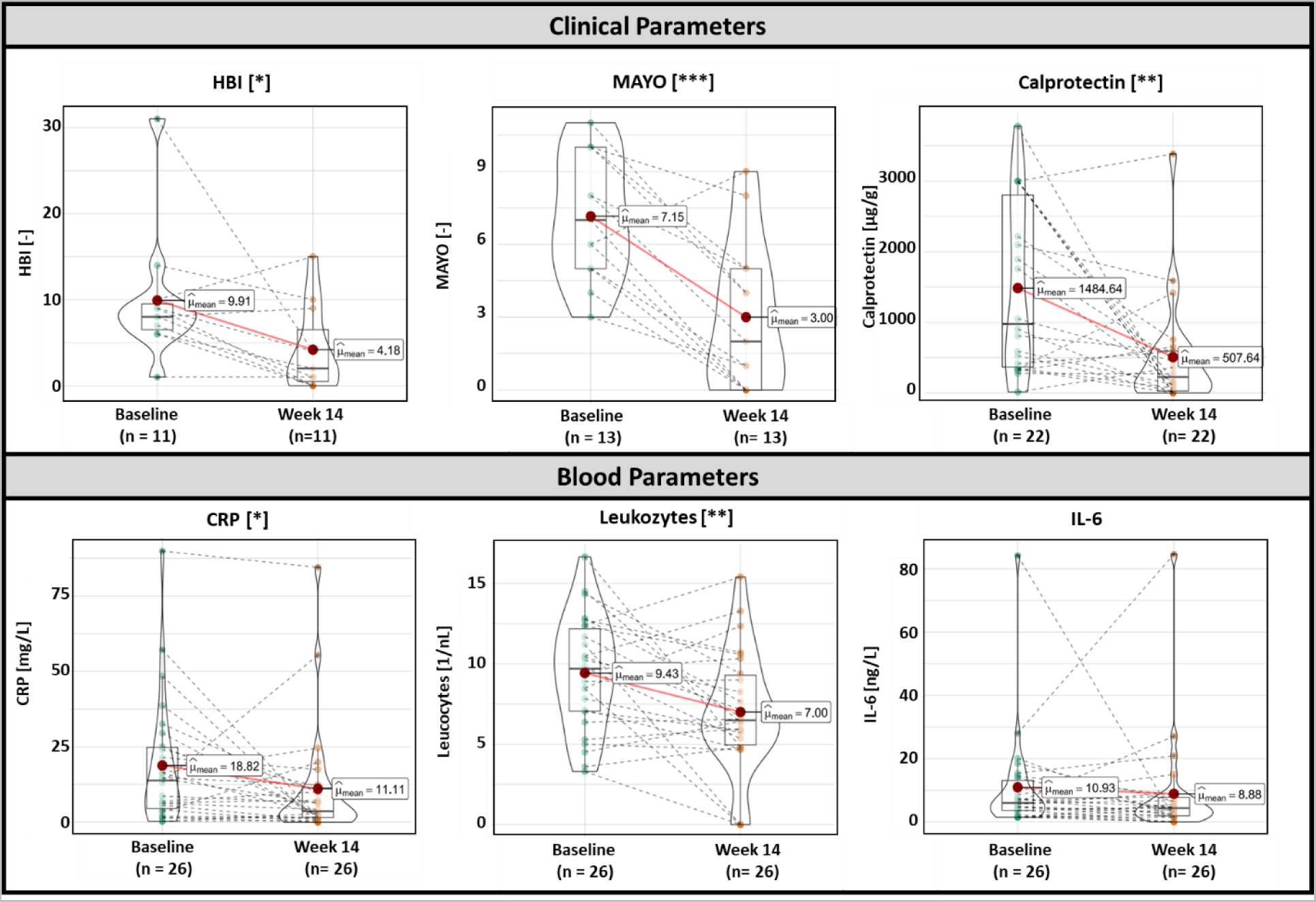
Clinical and blood parameters of individuals with IBD at baseline and after 14 weeks of therapy with Infliximab, Vedolizumab, or Olamkicept. Data are presented as violin plots, displaying the distribution of the data. Boxes depict the inner quartile range, with the median (black line) and mean (red dot). The outer shape clarifies the kernel probability density of the data. The dashed black lines indicate changes in the values for each patient. To analyze differences in paired samples Wilcoxon signed-rank test was used (* p<0.05, ** p<0.01, *** p<0.001). Abbreviations: HBI (Harvey Bradshaw Index), CRP (C-reactive protein), IL-6 (Interleukin-6)

### Metaproteomics analysis

To more specifically characterize the developments of the host inflammation and metabolism as well as the gut microbiome due to the induction of remission, we compared the fecal metaproteomes of remitting patients (n=12) at baseline (BL), and after 14 weeks (W14) of biologics therapy. Furthermore, we analyzed the changes in the fecal metaproteomes from patients (n=12) who did not achieve remission throughout the therapy and, to check for possible metaprotein signatures predictive of therapy success, we compared the metaproteomes of the two groups of patients before the therapy started.

On average over all measured samples, 10,268 (±2,149) metaproteins were identified and 31 % of spectra were assigned to bacteria, 1 % to archaeal species, and 0.5 % were assigned to viral proteins. Moreover, 40 % of spectra were assigned to eukaryotic proteins (Supplementary Table 1).

### Fecal metaproteome alterations through therapy in patients with remission

Numerous changes in the fecal metaproteomes of remitting patients (n=12) were observed. The proportion of spectra assigned to human proteins decreased significantly from 15.7 % to 12.8% (p <0.01) after the therapy in remitting patients (Figure 2A). Furthermore, 1,164 proteins were identified with significantly altered abundance (p-value<0.05, Figure 2B). These included human proteins such as immunoglobulin lambda variable 1-44 (p<0.001, fold change W14/BL: 0.52), lactotransferrin (p<0.001, fold change W14/BL: 0.3), progranulin (p<0.001, fold change W14/BL: 0.43), stomatin-like protein 3 (p<0.001, fold change W14/BL: 0.4) or peptidoglycan recognition protein 1 (PGLYRP1, p<0.001, fold change W14/BL: 0.42), which were significantly reduced. In addition, the abundance of various microbial proteins was also altered. The proteins with the lowest p-value included 50S ribosomal protein L7/L12 (p<0.001, fold change W14/BL: 0.31), 3-octaprenyl-4-hydroxybenzoate carboxy-lyase (p<0.001, fold change W14/BL: 0.60), DNA-directed RNA polymerase subunit beta (p<0.001, fold change W14/BL: 0.41), PAN2-PAN3 deadenylation complex catalytic subunit (p<0.001, fold change W14/BL: 0.33) and small ribosomal subunit biogenesis GTPase RsgA (p<0.001, fold change W14/BL: 0.32)(Table 1).

**Figure 2.**
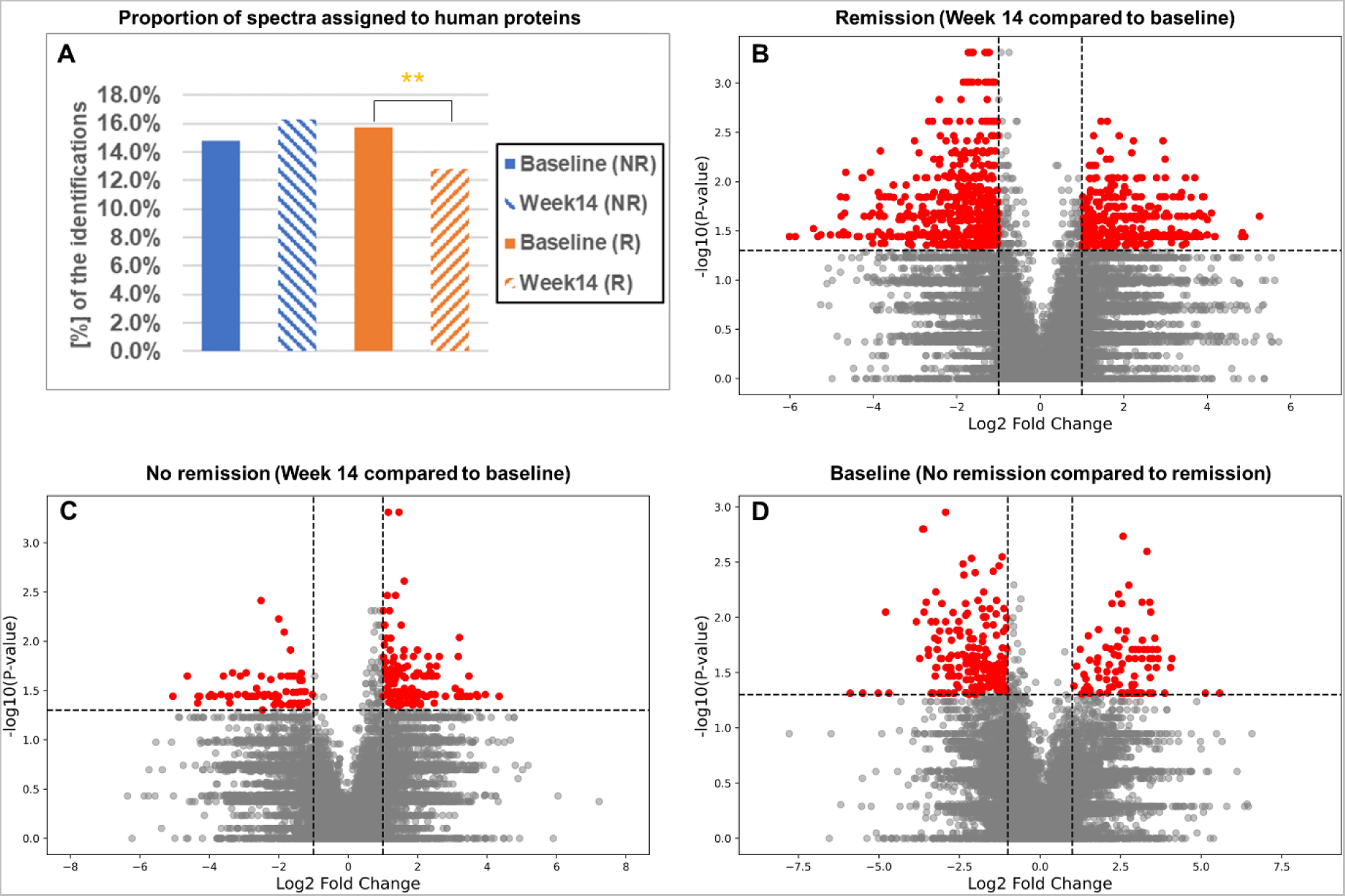
Proportion of spectra assigned to human proteins and differentially expressed metaproteins. A: Proportion of spectra assigned to human proteins in samples from non-remitting and remitting patients at baseline and after 14 weeks of therapy (Wilcoxon signed-rank test: ** *p* < 0.01. B: Volcano plot showing enrichment and depletion of metaproteins in baseline samples from non-remitting patients compared to baseline samples from remitting patients. C: Volcano plot showing enrichment and depletion of metaproteins in samples from remitting patients after 14 weeks of therapy compared to baseline. D: Volcano plot showing enrichment and depletion of metaproteins in samples from non-remitting patients after 14 weeks of therapy compared to baseline. In (B), (C), and (D), red dots describe sequences that have been enriched or depleted by factor 2 with a significance level of p < 0.05, calculated by Wilcoxon signed-rank test (B, C) and Wilcoxon rank sum test (D). The p-values and fold changes depicted in (B), (C), and (D) are shown in Table S2.

**Table 1.**
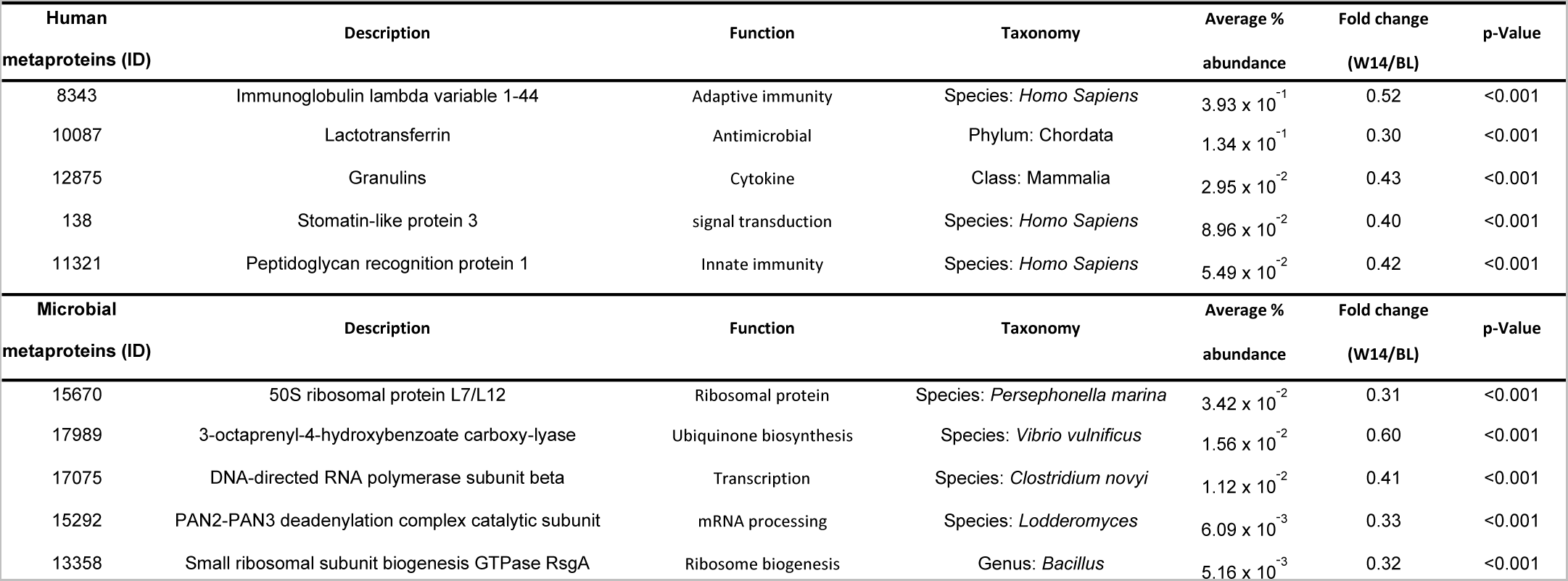
Significantly altered human and microbial metaproteins with lowest p-values (calculated with Wilcoxon signed-rank test) in samples from patients with remission at week 14 compared to baseline. Unknown metaproteins were excluded. The taxonomy column shows the lowest confirmed taxonomic rank. Metaproteins assigned to the kingdom Metazoa were considered as host (human) metaprotein. Wilcoxon rank sum test was used to analyze differences between unpaired samples (n=24). The function was inferred by UniProt keywords or GO term annotation.

| Human<br>metaproteins (ID) | Description | Function | Taxonomy | Average %<br>abundance | Fold change<br>(W14/BL) | p-Value |
| --- | --- | --- | --- | --- | --- | --- |
| 8343 | Immunoglobulin lambda variable 1-44 | Adaptive immunity | Species: *Homo Sapiens* | $3.93 \times 10^{-1}$ | 0.52 | <0.001 |
| 10087 | Lactotransferrin | Antimicrobial | Phylum: Chordata | $1.34 \times 10^{-1}$ | 0.30 | <0.001 |
| 12875 | Granulins | Cytokine | Class: Mammalia | $2.95 \times 10^{-2}$ | 0.43 | <0.001 |
| 138 | Stomatin-like protein 3 | signal transduction | Species: *Homo Sapiens* | $8.96 \times 10^{-2}$ | 0.40 | <0.001 |
| 11321 | Peptidoglycan recognition protein 1 | Innate immunity | Species: *Homo Sapiens* | $5.49 \times 10^{-2}$ | 0.42 | <0.001 |
| Microbial<br>metaproteins (ID) | Description | Function | Taxonomy | Average %<br>abundance | Fold change<br>(W14/BL) | p-Value |
| 15670 | 50S ribosomal protein L7/L12 | Ribosomal protein | Species: *Persephonella marina* | $3.42 \times 10^{-2}$ | 0.31 | <0.001 |
| 17989 | 3-octaprenyl-4-hydroxybenzoate carboxy-lyase | Ubiquinone biosynthesis | Species: *Vibrio vulnificus* | $1.56 \times 10^{-2}$ | 0.60 | <0.001 |
| 17075 | DNA-directed RNA polymerase subunit beta | Transcription | Species: *Clostridium novyi* | $1.12 \times 10^{-2}$ | 0.41 | <0.001 |
| 15292 | PAN2-PAN3 deadenylation complex catalytic subunit | mRNA processing | Species: *Lodderomyces* | $6.09 \times 10^{-3}$ | 0.33 | <0.001 |
| 13358 | Small ribosomal subunit biogenesis GTPase RsgA | Ribosome biogenesis | Genus: *Bacillus* | $5.16 \times 10^{-3}$ | 0.32 | <0.001 |

The comparison of functionally grouped proteins showed that intestinal barrier (p=0.034, fold change W14/BL: 0.79), neutrophil granulocytes (p=0.009, fold change W14/BL: 0.56), "other immune cells" (p=0.003, fold change W14/BL: 0.51) and immunoglobulin light chain proteins (p<0.001, fold change W14/BL: 0.71) were significantly reduced in remission patients at week 14, compared to baseline (Figure 2 A, B). In contrast, we observed several microbial processes such as butyrate fermentation (p=0.042, fold change W14/BL: 2.76), the pentose phosphate pathway (PPW, p=0.007, fold change W14/BL: 1.79), succinate synthesis (p=0.016, fold change W14/BL: 1.71) and sugar (p=0.003, fold change W14/BL: 1.97) and peptide transport (p=0.029, fold change W14/BL: 2.83), that were significantly increased in patients with clinical remissions (Figure 1C). Furthermore, we observed changes in the abundance of proteins of the blood (p=0.052, fold change W14/BL: 0.33), microbial lactate fermentation (p=0.569, fold change W14/BL: 8.41) or the complement system (p=0.107, fold change W14/BL: 0.30) which however, were not significant.

### Fecal metaproteome alterations through therapy in patients without remission

In patients without remission (n=12), the proportion of spectra assigned to human proteins increased non-significantly from 14.8 % to 16.3 % and only 375 metaproteins changed significantly in abundance (p<0.05). Human metaproteins which increased significantly, include LAMP1 (p=0.002, fold change W14/BL: 3.05), polymeric immunoglobulin receptor (PIGR, p=0.005, fold change W14/BL: 1.58), pantetheinase (VNN1, p=0.005, fold change W14/BL: 1.70), calcium-activated chloride channel regulator 1 (CLCA1, p=0.007, fold change W14/BL: 1.84) and neutrophil defensin 3 (DEF3A, p=0.007, fold change W14/BL: 2.88). Microbial metaproteins which were significantly altered include e.g., ribosomal RNA large subunit methyltransferase H (p<0.001, fold change W14/BL: 2.76) and anthranilate synthase component 2 (p=0.003, fold change W14/BL: 2.56) (Table 2).

**Table 2.** Significantly altered human and microbial metaproteins with lowest p-values (calculated with Wilcoxon signed-rank test) in samples from patients without remission at week 14 compared to baseline. Unknown metaproteins were excluded. The taxonomy column shows the lowest confirmed taxonomic rank. Metaproteins assigned to the kingdom Metazoa were considered as host (human) metaprotein. The function was inferred by UniProt keywords or GO term annotation.

| Human metaproteins (ID) | Description | Function | Taxonomy | Average % abundance | Fold change (W14/BL) | p-Value |
| --- | --- | --- | --- | --- | --- | --- |
| 12709 | Lysosome-associated membrane glycoprotein 1 | host-virus interaction | Class: Mammalia | $9.25 \times 10^{-3}$ | 3.05 | 0.002 |
| 8549 | Polymeric immunoglobulin receptor | polymeric Ig binding | Class: Mammalia | $5.75 \times 10^{-1}$ | 1.58 | 0.005 |
| 11174 | Pantetheinase | acute inflammatory response | Class: Mammalia | $4.69 \times 10^{-2}$ | 1.70 | 0.005 |
| 102 | Calcium-activated chloride channel regulator 1 | calcium transport | Species: *Homo sapiens* | $3.80 \times 10^{-1}$ | 1.84 | 0.007 |
| 368 | Neutrophil defensin 3 | Antimicrobial | Order: Primates | $1.29 \times 10^{-1}$ | 2.88 | 0.007 |
| Microbial metaproteins (ID) | Description | Function | Taxonomy | Average % abundance | Fold change (W14/BL) | p-Value |
| 14309 | Ribosomal RNA large subunit methyltransferase | rRNA processing | Species: *Clostridium difficile* | $7.77 \times 10^{-3}$ | 2.76 | <0.001 |
| 15465 | Anthranilate synthase component 2 | tryptophan biosynthesis | Species: *Salmonella enterica* | $8.41 \times 10^{-3}$ | 2.56 | 0.003 |
| 11588 | Uncharacterized protein HI_0392 | acyltransferase activity | Species: *Bacillus subtilis* | $9.08 \times 10^{-3}$ | 2.01 | 0.005 |
| 11586 | Alpha-monoglucosyldiacylglycerol synthase | Carbohydrate metabolism | Species: *Streptococcus* | $1.03 \times 10^{-2}$ | 2.27 | 0.005 |
| 22126 | Putative oligopeptide transport ATP-binding protein | Translocase | Species: *Bacillus subtilis* | $8.44 \times 10^{-3}$ | 2.06 | 0.007 |

Even in patients who did not achieve remission, differences in the metabolic processes and host protein abundance in stool metaproteome were observed after 14 weeks of therapy (Figure 3). Proteins of the intestinal barrier (p=0.035, fold change W14/BL: 1.40), IG-J chains (p=0.017, fold change W14/BL: 2.22) and human carbohydrate metabolism (p=0.011, fold change W14/BL: 1.49) were significantly increased in abundance. An insignificant increase was observed for e.g., immunoglobulin light chains (p=0.850, fold change W14/BL: 1.07) and proteins from neutrophilic granulocytes (p=0.428, fold change W14/BL: 1.14). In contrast, the microbial metabolic pathways of formate fermentation (p=0.013, fold change W14/BL: 0.34) and pyruvate synthesis (p=0.004, fold change W14/BL: 0.50) were significantly decreased, whereas vitamin B12 transport (p<0.001, fold change W14/BL: 1.60) was significantly increased. Human blood (p=0.296, fold change W14/BL: 0.50) and complement system (p=0.266, fold change W14/BL: 0.65) proteins were not significantly decreased.

**Figure 3.**
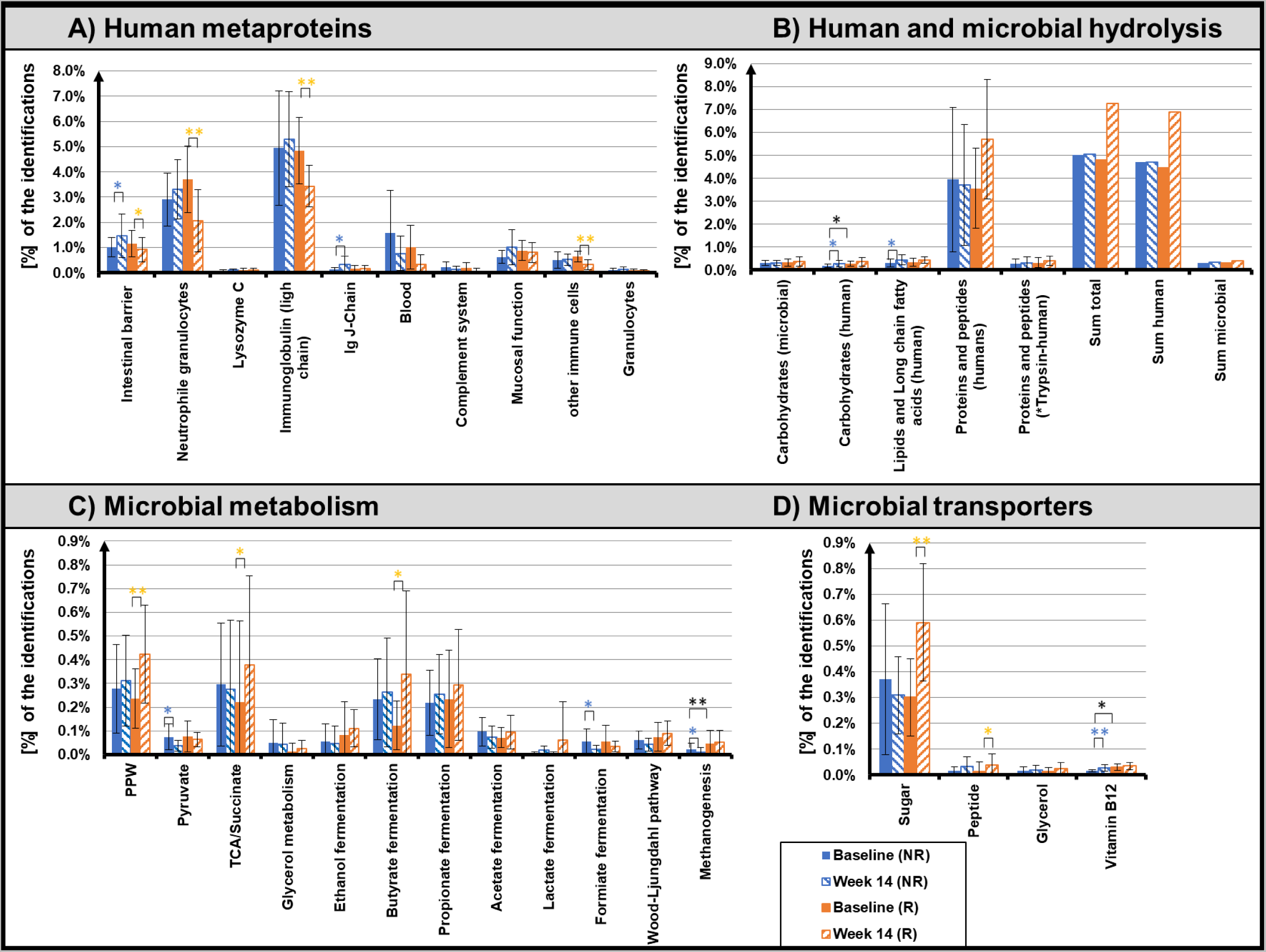
Overview of the functional assignment of identified metaproteins from patients at baseline and after 14 weeks of therapy. Data from patients who did not achieve remission are displayed in blue, data from patients who achieved remission are displayed in orange. Significant changes are marked with *. Identified metaproteins were functionally assigned to host metaproteins (A), microbial and human hydrolysis enzymes (B), microbial metabolism (C), and microbial transporters (D) (Supplementary Table 3). Data are presented as normalized average spectral abundance. All metaproteins assigned to the kingdom Metazoa were considered as host metaproteins. “Human trypsin” was shown as separated bar in (B) since its abundance was potentially increased due to the trypsin used for tryptic digestion. The Wilcoxon rank sum test was used to analyze differences in independent samples (n = 26, * p < 0.05, ** p < 0.01). The Wilcoxon signed rank test was used to analyze differences in dependent samples (n = 26, * p < 0.05, ** p < 0.01 for patients who achieved remission, n = 26, * p < 0.05, ** p < 0.01 for patients who did not achieve remission). Bar graphs show ± standard deviation.

### Fecal metaproteome alterations between patients with and without remission at baseline

Whereas the proportion of spectra assigned to human proteins was not significantly different at baseline between remitting and non-remitting patients (Figure 2A), a total of 550 metaproteins were significantly altered between patients with and without remission at baseline (p-value < 0.05, Figure 2B). Human proteins that were significantly increased in remission patients included DEF3A (p=0.003, ratio R/NR: 4.37), HLA class II histocompatibility antigen, DR alpha chain (p=0.003, ratio R/NR: 5.27), or CLCA1 (p=0.004, ratio R/NR: 2.74) (Table 3). In contrast, human metaproteins like centromere protein F were significantly increased in non-remitting patients (p=0.002, R/NR: 0.17). Significantly changed microbial proteins included riboflavin biosynthesis protein RibBA (p=0.002, ratio R/NR: 12.24), chloramphenicol acetyltransferase 2 (p=0.002, R/NR: 0.17) and polyphosphate kinase (p=0.001, only in patients with Remission).

**Table 3.** Significantly altered human and microbial metaproteins between patients with and without remission at baseline with lowest p-Values. Unknown metaproteins were excluded. The taxonomy column shows the lowest confirmed taxonomic rank. Metaproteins assigned to the kingdom Metazoa were considered as host (human) metaprotein. Wilcoxon rank sum test was used to analyze differences between unpaired samples (n=24). The function was inferred by UniProt keywords or GO term annotation.

| Human metaproteins (ID) | Description | Function | Taxonomy | Average % abundance | Fold change (R/NR) | p-Value |
| --- | --- | --- | --- | --- | --- | --- |
| 9938 | Double homeobox protein 4-like protein 4 | transcription regulation | Species: *Homo Sapiens* | $9.1 \times 10^{-2}$ | 7.65 | 0.001 |
| 23146 | Centromere protein F | cell cycle | Species: *Homo Sapiens* | $1.43 \times 10^{-3}$ | 0.17 | 0.002 |
| 368 | Neutrophil defensin 3 | antimicrobial | Order: Primates | $1.79 \times 10^{-1}$ | 4.37 | 0.003 |
| 17966 | HLA class II histocompatibility antigen, DR alpha chain | adaptive immunity | Species: *Homo Sapiens* | $8.36 \times 10^{-3}$ | 5.27 | 0.003 |
| 17342 | Calcium-activated chloride channel regulator 1 | calcium transport | Class: Mammalia | $1.29 \times 10^{-2}$ | 2.74 | 0.004 |
| Microbial metaproteins (ID) | Description | Function | Taxonomy | Average % abundance | Fold change (R/NR) | p-Value |
| 8869 | Riboflavin biosynthesis protein RibBA | riboflavin biosynthesis | Superkingdom: Bacteria | $1.65 \times 10^{-3}$ | 12.24 | 0.002 |
| 23145 | Chloramphenicol acetyltransferase 2 | antibiotic resistance | Species: *Escherichia coli* | $1.43 \times 10^{-3}$ | 0.17 | 0.002 |
| 23148 | UDP-N-acetylmuramoyl-tripeptide--D-alanyl-D-alanine | cell wall biogenesis/degradation | Species: *Synechocystis sp. PCC 6803* | $1.04 \times 10^{-2}$ | 0.10 | 0.003 |
| 20508 | Polyphosphate kinase | kinase | Class: Gammaproteobacteria | $2.03 \times 10^{-1}$ | only in patients | 0.003 |
| 3908 | N-acetylneuraminate epimerase | carbohydrate metabolism | Family: *Enterobacteriaceae* | $1.11 \times 10^{-3}$ | only in patients | 0.003 |

To disentangle host and microbial processes, we independently assessed metaproteomes attributed to humans or microbiota (Figure 3). Concerning the functional processes of the gut microbiome, the abundance of proteins involved in vitamin 12 transport was significantly higher (p=0.014, ratio R/NR: 1.86) and the abundance of proteins of microbial methanogenesis (p=0.003, ratio R/NR: 0.34) was found significantly lower in patients who achieved remission. (Figure 3C). Other non-significantly different processes in terms of their protein abundance included bacteriocin transport (p=0.059, ratio R/NR: 3.88), glycerol metabolism (p=0.159, ratio R/NR: 0.26) or succinate metabolism (p=0.143, ratio R/NR 0.74).

No processes to which human proteins were grouped, were significantly altered, except for proteins assigned to carbohydrate hydrolysis (p=0.024, ratio R/NR:). Proteins of neutrophil granulocytes (p=0.143, ratio R/NR: 1.28) and proteins of mucosal functions (p=0.060, ratio R/NR: 1.42) were present in lower abundance in patients who did not achieve remission.

### Identification of disease-specific biomarker candidates for monitoring the success of IBD therapy

For the identification of disease-specific biomarker candidates for monitoring or predicting the success of IBD therapy, only metaproteins with an average relative abundance of at least 0.01% in at least one patient group (R-BL, R-W14, NR-BL, NR-W14) were considered. The most promising human and microbial biomarkers, in our opinion, for which a pathogenesis contribution can be traced, are shown in Figure 4. The first microbial biomarker shown, anthranilate synthase component 2 (Meta-Protein 15465, *Salmonella enterica*) increases significantly during therapy in patients who did not achieve remission (NR-BL/NR-W14: p<0.005). Furthermore, 3-octaprenyl-4-hydroxybenzoate carboxy-lyase (Meta-Protein 17989, *Vibrio vulnificus*, R-BL/R-W14: p<0.001) was found significantly decreased and glutamate dehydrogenase (Meta-Protein 1031, Unknown Superkingdom, R-BL/R-W14: p=0.002) was found significantly increased in remitting patients.

**Figure 4.**
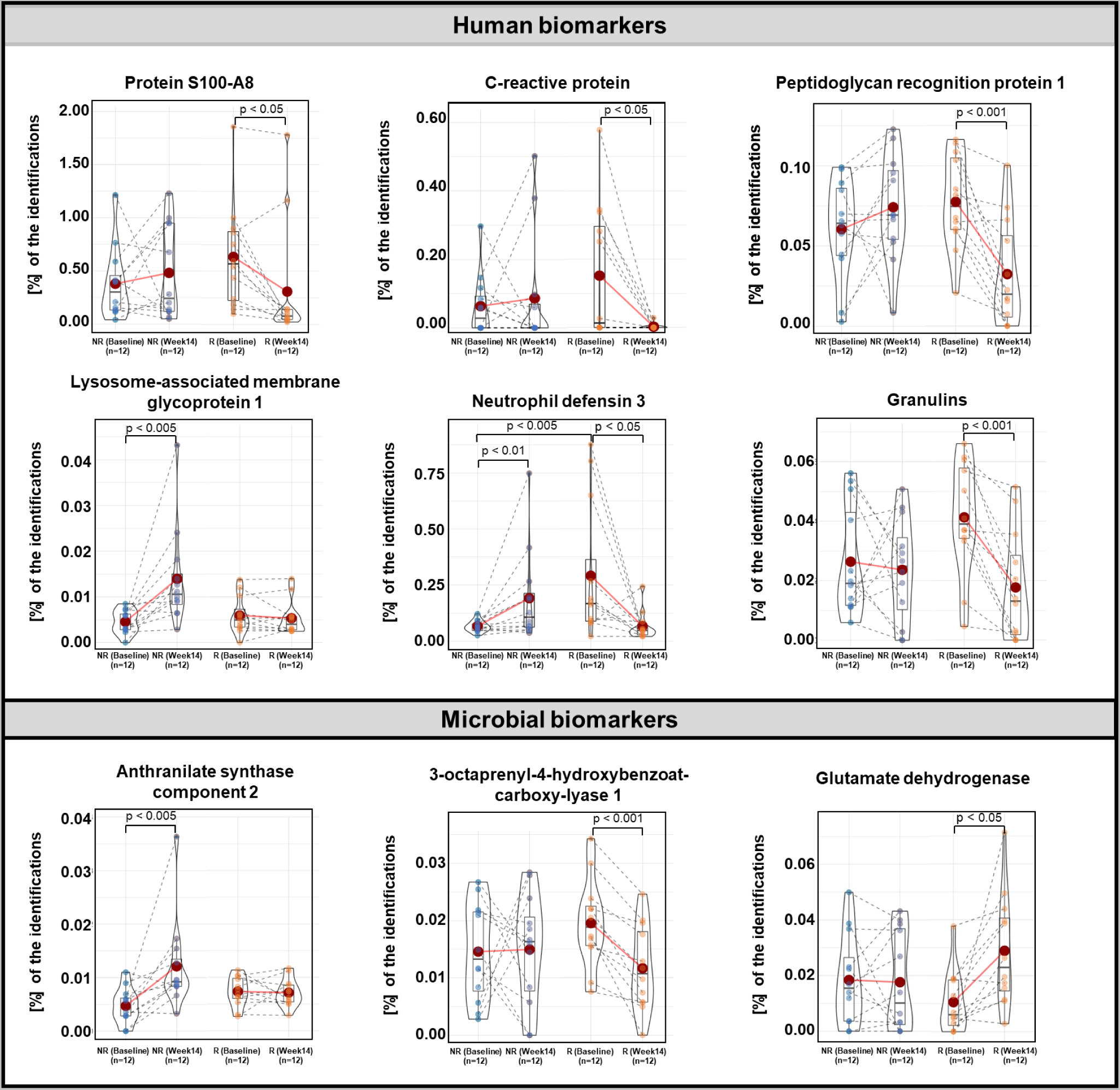
Panel of potential marker metaproteins as violin plots. Relative abundance of the marker metaproteins in fecal samples from IBD patients who achieved remission (R) and IBD patients who did not achieve remission (NR) at baseline and after 14 weeks of biologics therapy. The means are illustrated as red dots. P-values were calculated with Wilcoxon signed-rank test (between timepoints of remitting or non-remitting patients) and Wilcoxon rank sum test (between remitting and non-remitting patients at baseline).

The abundance of the human proteins S100-A8 (R-BL/R-W14: p=0.012) and CRP (R-BL/R-W14: p=0.036) decreased in patients with remission in the course of therapy. Significant changes were also observed for the human protein DEF3A (R-BL/NR-BL: p=0.001, R-BL/R-W14: p=0.021, NR-BL/NR-W14: p=0.007). Only in patients without remission does the human protein LAMP1 (NR-BL/NR-W14: p=0.003) increase. In contrast, the PGLYRP1 decreases significantly in patients with remission (R-BL/R-W14: p=0.001). Likewise, the protein Granulins decreases significantly in remission patients (R-BL/R-W14: p<0.001).

In a first attempt to validate the finding from the metaproteomic biomarker discovery, LAMP1, as a probable indicator for the failure of the therapy, was measured with ELISAs in fecal washes of UC patients (Figure S1). A large but statistically insignificant (p=0.069) increase in the abundance of LAMP1 in was found in patients, who were not in remission.

## 4. Discussion

### Fecal metaproteome alterations through therapy in patients with remission

With the help of the analysis of the fecal metaproteomes from patients who achieved remission, a large number of changes in the course of therapy could be determined. These are obvious by the large number of significantly altered proteins and the significantly decreased proportion spectra assigned to human proteins in the course of the therapy, which was described previously to be indicative of disease activity [31].

The changes indicate on the one hand that the immune response of this group of patients has decreased and on the other hand that proteins of the microbial metabolism are present in increased abundance.

The significantly decreased abundance of intestinal barrier proteins, as well as the decrease in blood proteins, indicate an increased integrity of the epithelial layer of the intestine [32, 33]. Furthermore, the decreased abundance of proteins from neutrophilic granulocytes is a sign of less translocation to the gut of these immune cells [34, 35], which are an important driver of inflammation in IBD. The significantly decreased human proteins, immunoglobulins, lactotransferrin, or progranulin have already been associated with IBD [36–38]. Their decline in abundance suggests an ameliorated disease course. The significant decrease in PGLYRP1 also indicates less dysbiosis with the gut microbiome, as this protein, which is also part of neutrophil extracellular traps (NETs), recognizes bacterial cell wall structures and initiates the inflammatory response [39, 40].

The reduced dysbiosis is also recognizable in the significantly increased abundance of microbial metabolic pathways, such as the PPW, the tricarboxylic acid cycle /succinate metabolism, or butyrate fermentation. The PPW is crucial for biomass increase and growth and an increased abundance of its enzymes may indicate a prosperous intestinal microbiome [41], further supported by increased transport of sugars and peptides. The increased metabolism of succinate, which promotes the release of pro-inflammatory cytokines, could prevent the accumulation of it in the feces, which is a phenomenon observed in IBD inflammation, and thereby further improve growth conditions [42]. Finally, butyrate fermentation in particular is of great importance for the well-being of the patient, as butyrate serves as an energy source for colonocytes and has great anti-inflammatory potential [43, 44]. The increased synthesis of butyrate can also serve as an indicator of the stability of remission [45].

### Fecal metaproteome alterations through therapy in patients without remission

Based on the altered processes and proteins of the patients who did not achieve remission, a strongly deviating reaction of the intestinal immune system and the gut microbiome to the therapy can be stated. Immune-relevant processes and protein groups, like proteins from neutrophils or immunoglobulins, which showed a clear decline in patients with remission, are not significantly decreased in this group of patients. The increase of immunoglobulin light chain and immunoglobulin J-chain could also be a hint at the formation of anti-drug antibodies, which are a major cause of therapy failure in biologics treatment [46], The increased formation of microbiome targeting immunoglobulins was observed in active IBD [47], so th95e target of those antibodies needs to be elucidated. Nevertheless, the secretion of immunoglobulins into the gut lumen depends on the PIGR [48], which is one of the most significantly increased proteins in non-remitting patients, supporting the hypothesis of an increased antibody response.

Other individual proteins, that have been described as the IBD susceptibility locus, such as VNN1 [49], or that are associated with the immune response in IBD, like LAMP1 [50], are present in significantly increased abundance after therapy. These findings indicate an ongoing inflammation whose character has changed due to the induction of therapy. The reason for an ongoing inflammation could be the expansion of apoptosis-resistant immune cells, which was observed in non-responding IBD patients in reaction to biologics therapy. Immune modulatory therapies have also been shown to increase LAMP1 abundance [51], which is often used to measure degranulation and cytotoxic potential of cytotoxic lymphocytes [52]. Increased cytotoxicity was observed in IBD patients in association with increased apoptosis in the intestinal mucosa [53]. Identification of epithelial proteins like VNN1, PIGR, and CLCA1 among the most significantly increased proteins and the significant increase in the abundance of intestinal barrier proteins in general, would support this hypothesis. Increased abundance of proteins from neutrophilic granulocytes, especially antimicrobial DEF3A, could be a reaction to increased microbial evasion due to barrier defects. As microbial metabolite synthesis, like butyrate fermentation or formate synthesis, which is significantly decreased in non-remitting patients, potentially limits the cytotoxic effects of immune cells [54, 55], the lack of a healthy microbiome could contribute to therapy failure.

Nevertheless, there are some changes in the metaproteome of these patients that indicate at least a slightly beneficial response to therapy. Proteins of microbial vitamin B12 transport were significantly increased, which could indicate a reduced dysbiosis of the gut microbiome [56]. Furthermore, the abundance of human blood proteins was halved. As fecal hemoglobin was previously identified as a biomarker for intestinal inflammation [57], the decline of blood proteins could indicate at least a minor amelioration of the disease. This is also supported by the reduction of proteins coupled to the complement system. As this pathway is part of the innate immune response and the response to microbial-associated molecular patterns this could also be a hint to less pronounced microbial dysbiosis.

### Fecal metaproteome alterations between patients with and without remission at baseline

As there are no reliable biomarkers to predict the success of a therapy and clinicians often empirically try different drugs to achieve remission [58], our final comparison aimed to identify metaprotein patterns, that are likely to predict the success of biologics therapy before the therapy started.

In healthy individuals, the defense against pathogenic organisms in the human intestine takes place through a balanced reaction of the innate and adaptive immune system [59]. Increased immune response is seen at the onset of therapy, particularly in patients who subsequently achieve remission. Proteins of the innate immune system, such as neutrophil DEF3A, and the adaptive immune system, such as HLA class II histocompatibility antigen, are increased significantly in remitting patients. A high amount of alpha-defensins was indeed described as a prediction of therapy response in IBD therapy before [60].

Furthermore, ROS are an important instrument of the intestinal immune response. But they represent a double-edged sword in the pathogenesis of IBD, as a fine-tuned regulation is necessary to eliminate pathogens, but at the same time avoid killing commensal microorganisms and damage tissue. The increased abundance of microbial proteins that synthesize the antioxidants riboflavin and polyphosphate in patients, who achieve remission suggests an active contribution of the gut microbiome to tissue protection in these individuals [61]. In contrast, microbial processes that act as a sink for the anti-inflammatory molecule hydrogen [62], such as methanogenesis and glycerol metabolism, are more prevalent in patients who do not achieve remission. These changes indicate an imbalance of the immune response and the microbiome concerning oxidative stress, which possibly promotes the depletion of beneficial microorganisms and tissue damage, which seems to extend during the therapy, as stated above. This hypothesis is underlined by the increased abundance of blood proteins in patients without remission. Furthermore, the increased abundance of chloramphenicol acetyltransferase 2 in non-remitting patients hints at an involvement of the gut resistome, which is expanded in many diseases [63].

For other significantly changed proteins like Double homeobox protein 4-like protein 4, a transcription factor or centromere protein F, a marker for poor prognosis in cancer [64, 65], no connection to IBD or gut immunity is described underlining the knowledge gaps still existing.

In general, predictive biomarkers are described as dependent on used biologic [66], which renders the general assessment of predictive biomarkers hard in the present study. Nevertheless, as microbiome fitness and human protein biomarkers are included in nearly all predictive models, the prediction of therapy success using metaproteomics appears feasible. Taking into account other omics levels, like metabolomics, and a personalized approach, thereby benefitting from knowledge about e.g., individual polymorphisms of a patient, would further improve prediction potential [67].

### Identification of disease-specific biomarkers

To be able to predict the success of therapy before medication or to assess the effectiveness of ongoing therapy, the search for biomarkers is a central topic of ongoing research. As we opted to identify markers, which are present in easily detectable abundances, we only considered metaproteins with an average relative abundance of at least 0.01% in at least one patient group (R-BL, R-W14, NR-BL, NR-W14).

In particular, fecal calprotectin (FC) and CRP have been investigated in a large number of studies using antibody-based methods such as ELISA [68–70]. Although their use is often hindered by disadvantages such as a lack of specificity [71], we considered them as biomarkers in our cohort. Both show a significant decrease in patients with remission, but there is no significant difference between patients with and without remission at baseline to predict therapy success. Taken together with the observation of outlying individuals, the need for more specific biomarkers or a panel of biomarkers is obvious.

The earlier-mentioned increased antimicrobial potential in patients without remission is supported by the consideration of DEF3A. Furthermore, the data presented in Figure 4 regarding the abundance of the proteins of neutrophil granulocytes are depicted reliably, so the protein seems to be suitable as a marker protein for the invasion of this type of leukocyte. The violin plot (Figure 4) further implies that a high amount of DEF3A, which has broad antimicrobial activity [72], in a patient’s feces predicts therapy success at baseline, whereas a low amount could be proof of a successful therapy after termination. Interestingly, high DEF3A was also evaluated as a positive predictive marker in cancer immunotherapy [73].

A marker for reduced dysbiosis in remission patients could also be the human PGLYRP1. Since this protein is involved in antigen recognition [40], the decrease in this protein may indicate reduced invasion of intestinal tissue by microorganisms. The significant decrease in granulins also indicates a decrease in immune response and inflammation in remission patients.

In contrast, the significant increase in LAMP1, an essential protein of natural killer cell cytotoxicity [74], may indicate a change in the nature of inflammation in patients without remission. This means physicians may have to change the therapeutic strategy, for example to antibodies against NKG2D, which induces cytolytic natural killer cells and T-cells upon interaction with stress-related molecules [75]. The validation of LAMP1 with an ELISA also showed a higher abundance in non-remitting patients, but the increase was not significant. Nevertheless, the strong elevation in distinct patients showed the relevance of the protein for specific individuals supporting a precision medicine approach to the treatment of IBD patients.

As all human markers depicted in Figure 4 are also described in other malignancies [73, 76–80], their specificity in IBD has to be evaluated further. Since microbial dysbiosis and associated functional and taxonomic differences are hallmarks of IBD, there are also many attempts to discover microbial biomarkers for disease monitoring or responsiveness to therapies. The significant differences in the microbial biomarkers proposed in Figure 4 show, that they are suitable for disease monitoring. Markers for the different development of microbial metabolism in patients with and without remission could be anthranilate synthase component 2 (Meta-Protein 15465, *Salmonella enterica*), which is involved in of the tryptophan metabolism and quorum sensing [81]. An increased tryptophan metabolism is under investigation to aggravate disease activity in IBD patients [82]. Furthermore, anthranilate has antimicrobial potential and by increasing in patients who did not achieve remission, the antimicrobial milieu in these patients could be further worsened. Furthermore, the species the metaprotein is annotated to, *Salmonella enterica*, is described to benefit from inflammation, to overcome colonization resistance and to play a role in the onset of IBD [83, 84].

In contrast a decreasing abundance of 3-octaprenyl-4-hydroxybenzoate carboxylyase (Meta-Protein 17989, *Vibrio vulnificus*), which catalyzes an intermediate step in the biosynthesis of ubiquinone, an antioxidant and tested as supplement in inflammatory diseases, could serve as a marker of less oxidative stress and a more hospitable environment for microorganisms. A similar sign of regrowth of a beneficial gut flora could be glutamate dehydrogenase (Meta-Protein 1031, Unknown Superkingdom), which has great importance for microbes in gut colonization and is the most abundant metaprotein found in fecal samples from healthy individuals [85, 86]. Nevertheless, as the gut microbiome underlies huge interindividual and intraindividual changes, finding a single microbial marker protein remains a challenging task, and a panel of marker proteins or the functional assessment of the microbiome might be more promising.

Despite the ability of metaproteomics to characterize the induced changes in the remission of IBD patients in contrast to non-remitting patients, the present study has limitations. Due to the small number of study participants, the power of the statistical data analysis is limited. Furthermore, the field of participants is quite heterogeneous with regard to their disease. In the evaluation, neither the type of disease (UC, CD) or its local occurrence nor the type of therapeutic agent was considered. Due to the different mechanisms of action of the drugs, different changes in the gastrointestinal microbiome are to be expected [87], leading to specific metaproteome patterns especially between patients treated with different biologics. This could minimalize the observations on the microbial aspects of the analysis. Larger clinical panels, taking into account the different biologics and disease subtypes, to evaluate the differences identified here would be useful.

Furthermore, the influence of environmental influences, e.g. changes in the patient’s diet in the course of therapy, on the metaproteome of the study participants is difficult to assess, which could make the already large interindividual differences even more significant.

Moreover, many identified meta-proteins, some of which changed significantly in the course of therapy, could not be annotated. Analyzing different metaproteomic data sets may hold potential to help in annotation of these potentially disease relevant features [88].

Fecal metaproteomics enabled us to monitor the success of biological-based IBD therapy deduced from proteins linked with inflammation and bacterial metabolisms, showing its potential for clinical application. Remitting patients showed a decrease in immune functions, in particular, decreased abundance of neutrophilic proteins and an increased abundance of several microbial functions, like butyrate synthesis. The fecal metaproteomes of patients with an unsuccessful therapy were marked by increased abundance of immunoglobulin proteins and signs of cytotoxicity and tissue damage. Furthermore, we proposed several new potential biomarker candidates, which could also be of interest for further antibody-based monitoring of IBD therapy.

## Data Availability

All data produced in the present study are available upon reasonable request to the authors.
The mass spectrometry proteomics data have been deposited to the ProteomeXchange Consortium via the PRIDE partner repository with the dataset identifier PXD053257.

## Abbreviations

CD: Crohn’s disease
CLCA1: Calcium-activator chloride channel regulator 1
CRP: C-reactive protein
DEF3A: Neutrophil defensin 3
E.C.: Enzyme comission
eMayo: Endoscopic Mayo
FASP: Filter-aided sample preparation
FC: Fecal calprotectin
HBI: Harvey-Bradshaw index
IBD: Inflammatory bowel disease
IL-6: Interleukin 6
KEGG: Kyoto Encyclopedia of Genes and Genomes
LAMP1: Lysosomal-associated membrane protein 1
NET: Neutrophil extracellular trap
PIGR: Polymeric immunoglobulin receptor
PPW: Pentose phosphate pathway
TCA: Tricarboxylic acid
UC: Ulceratice colitis
VNN1: Pantetheinase

## 6. Acknoledgements

Not applicable.

## 7. Data Availability Statement

The mass spectrometry proteomics data have been deposited to the ProteomeXchange Consortium via the PRIDE partner repository with the dataset identifier PXD053257.

Reviewer access details

Log in to the PRIDE website using the following details:

Project accession: PXD053257

Token: ZUI01Soinr2K

Alternatively, reviewer can access the dataset by logging in to the PRIDE website using the following account details:

Username

Password: z6IsKunGzcvs

## 9. Author contributions

- Conceptualization: R.H., K.A.
- Experiments metaproteomics: J.L., P.H.
- ELISA experiments: L.W., S.N., F.T., S.T.
- Data evaluation: J.L., M.W., L.W., S.N., F.T., S.T.
- Bioinformatics: K.S., J.L., M.W.
- Graphical abstract: K.S.
- Supervision: R.H., K.A.
- critical revision of the manuscript: D.B., M.G., R.H., K.A., K.S., U.R., P.R.
- writing—original draft: M.W.
- writing—review and editing: R.H., K.A.

All authors have read and agreed to the published version of the manuscript.

