## Supplementary Note 1 for "Fecal metaproteomics enables functional characterization of remission in patients with inflammatory bowel disease"

**Supplementary Note 1: Detailed description of the wet-lab methods used for metaproteome analyses**

The metaproteomic analyses were performed similarly to a previously published protocol [1]. Approximatly 200 µg of fecal samples dissovled in 200 µL 1% sodium dodecyl sulfate solution, 1 g silica beads, 400 μL 2 M sucrose solution and 700 μL phenol dissolved in water were shaken in a ball mill (MM400 Retsch, Düsseldorf, Germany) for 10 min at 30 Hz. After centrifugation for 10 min at 10,000  xg at room temperature (RT), the phenolic phase was transferred into a new reaction tube, mixed with the same volume of 1M sucrose solution and shaken in the ball mill for 5 min at 1800 rpm. After centrifugation (10,000 xg, RT, 10 min) the protein containing phenolic phase was transferred into a new reaction tube and the proteins were prcipitated at -20°C by the addition of ice-cold 100 mM ammonium acetate in methanol. After another centrifugation and precipitation step, the supernatant was removed. The pellet was dried and dissolved in7 M urea, 2 M thiourea (urea buffer) as well as 0.01 g/mL dithiothreitol**.**

The amidoblack assay was used for the quantification of protein content [2, 3]. 50 µL of samples were precipitated with 300 µL aof amidoblack solution (0.26 mg/L amido black, 90% methanol, 10% acetic acid) and centrifuged (16,400× g, RT, 5 min). After removal of the supernatant, the pellet was washed twice with 500 µL 10% acetc acid in methanol. The supernatant was decanted after a further centrifugation step (16,400× g, RT, 5 min) and the stained protein pellet was dissolved in 1 µL 0.1 M sodium hydroxide solution. The absorption was measured with a photometer (Spectrophotometer Genesys 10S UV-Vis, Thermo Scientific, Waltham, USA) at a wavelength of 615 nm and the protein concentration determined using a calibration curve with defined amounts of bovine serum albumin.

A modified version of the Filter-aided sample preparation (FASP) protocol was used for tryptic digestion [4]. Protein solution containg 10 µg of protein was filled up to 200 µL with 8 M urea and loaded on a FASP filter (Pall Nanosep, MWCO 10 kDa Omega, VWR, Dresden). The buffer was removed by centrifugation ( 10,000xg, RT, 10 min). The protein on the filter was washed with 200 µL urea buffer. To reduce the disulfide bonds, 100 µL 40 mM dithiothreitol was added and incubated for 20 min on a thermomixer (300 rmp) at 56°C. After centrifugation, 100 µL of 55 mM iodoacetamide was added. After incubation for 20 min in the dark and subsequent cenrifugation, the proteins were washed once with urea buffer, followed by three washing steps with 100 µL 50 mM ammonium bicarbonate. For trypsinization, 200 µL of trypsin in 50 mM ammonibicarbonate was added to achieve an enzym-subtrate ratio of 1:100 and incubated for 2 h at 37°C on a thermomixer (300 rpm). After a further centrifugation step, the remaining peptides on the filter were rinsed by adding 50 µL of 5% acetonitrile in 50 mM ammonium bicarbonate and 50 µL LC-MS grade water, respectivly. The peptide solutions were dried in a vaccum centrifuge and solubilized in 30 µL of 0.1% trifluoroacetic acid in LC-MS grade water.

The peptide solutions were analyzed using an UltiMate 3000 RSLC nano splitless liquid chromatography system (Thermo Fisher Scientific, Bremen, Germany), coupled online to a timsTOF™ pro mass spectrometer (Bruker Daltonik, GmbH, Bremen). After injection, the peptides were loaded isocratically on a trap column (Acclaim PepMap100 C18, Thermo Fisher Scientific, Bremen, Germany) for desalting and concentration. Chromatographic separation was performed on a Dionex Acclaim PepMap C18 RSLC nano reversed-phase column (Thermo Fisher Scientific, Bremen, Germany). A flow rate of 400 nL/min was applied using a binary A/B-solvent gradient (solvent A: 100 % LC-MS grade water, 0.1 % formic acid; solvent B: 100 % acetonitrile, 0.1% formic acid)) starting with 5% solvent B and increasing over a 120 min gradient to 35% solvent B.

A positive mass spectrometric measurement was used in a data-dependent MS/MS method. Parallel Accumulation Serial Fragmentation (PASEF) scan mode was executed using trapped ion mobility spectrometry (TIMS) in a range of 0.6 to 1.6 V*s/cm2 and 10 PASEF MS/MS scans per cycle. The MS/MS data acquisition was conducted over a mass range from 100 to 1700 m/z using collision-induced dissociation (CID) with active exclusion of the same precursor after 1 spectra for 24 seconds or a release if intensity/previous intensity exceeded 4. The resulted fragment ions were analyzed in a TOF analyzer.
